# Methylation profiles at birth linked to early childhood obesity

**DOI:** 10.1101/2024.01.12.24301172

**Authors:** Delphine Lariviere, Sarah J.C. Craig, Ian M. Paul, Emily E. Hohman, Jennifer S. Savage, Robert O. Wright, Francesca Chiaromonte, Kateryna D. Makova, Matthew L. Reimherr

## Abstract

Childhood obesity represents a significant global health concern and identifying risk factors is crucial for developing intervention programs. Many ‘omics’ factors associated with the risk of developing obesity have been identified, including genomic, microbiomic, and epigenomic factors. Here, using a sample of 48 infants, we investigated how the methylation profiles in cord blood and placenta at birth were associated with weight outcomes (specifically, conditional weight gain, body mass index, and weight-for-length ratio) at age six months. We characterized genome-wide DNA methylation profiles using the Illumina Infinium MethylationEpic chip, and incorporated information on child and maternal health, and various environmental factors into the analysis. We used regression analysis to identify genes with methylation profiles most predictive of infant weight outcomes, finding a total of 23 relevant genes in cord blood and 10 in placenta. Notably, in cord blood, the methylation profiles of three genes (*PLIN4, UBE2F,* and *PPP1R16B*) were associated with all three weight outcomes, which are also associated with weight outcomes in an independent cohort suggesting a strong relationship with weight trajectories in the first six months after birth. Additionally, we developed a Methylation Risk Score (MRS) that could be used to identify children most at risk for developing childhood obesity. While many of the genes identified by our analysis have been associated with weight-related traits (e.g., glucose metabolism, BMI, or hip-to-waist ratio) in previous genome-wide association and variant studies, our analysis implicated several others, whose involvement in the obesity phenotype should be evaluated in future functional investigations.

## Introduction

Obesity affects over 40% of Americans^1^, including nearly 20% of children^2^. Childhood obesity is associated with various disorders across the life course, including hypertension, hypercholesterolemia, and insulin resistance^3,4,5^. To maximize the benefit of preventive interventions^6,7,8^, early identification of children who are most at risk for developing obesity is paramount.

Weight is a complex trait influenced by many factors, including the environment (e.g., diet, activity level, medications), genetics, epigenetics, the microbiome, and the metabolome of individuals. Previous studies have indicated that 40-80% of variation in BMI can be explained by genetic factors^9,10^. However, the cumulative effect of single nucleotide polymorphisms (SNPs) identified so far does not account for all of the variation attributed to genetics. Specifically, earlier genome-wide association studies (GWASs) have only been able to explain approximately 3% of variation in BMI, and more recent studies^11^ considering SNPs significant at the genome-wide level explain up to 6% of such variation. Less stringent studies or meta-analyses raised this percentage to over 20% (reviewed in Bouchard *et al.* 2021^10^), but still failed to explain the observed heritability in obesity, which approaches 50%^10^. Despite this gap, polygenic risk scores (PRSs) are being widely developed to combine variants from GWASs to assess an individual’s risk for disease^12^. These scores have been developed for adults^13^ and more recently for children^14^.

In addition to genetic factors, epigenetic modifications could provide important insights into an individual’s risk for obesity because they can be heritable when located in the germline, and modifiable by environmental factors^15^. Epigenomics, the study of epigenetic modifications on a genome-wide scale, is a field of research that links genes and disease to provide a complex picture accounting for changes due to environmental influences across the lifetime. The most common epigenetic modification of DNA is cytosine methylation at CpG sites. Methylation plays a role in repressing gene expression when located in regulatory regions^16^ and has been linked to active gene transcription when located within the gene body^17^. The proposed molecular mechanisms of gene body methylation range from silencing of repetitive elements^18^ to affecting nucleosome positioning^19^ and histone modifications^20^. Analogous to constructing PRSs with SNP data, methylation risk scores (MRSs) have been recently developed^21,22^. MRSs are linear combinations of methylation states across multiple CpG sites and may be useful in the clinical setting as these epigenetic marks can be influenced by environmental conditions and thus could be used to monitor changes in disease risk over time^21^.

In the context of childhood obesity, some studies have shown differences in peripheral blood methylation profiles between children with and without overweight^23,24^. Other studies have identified CpG loci whose methylation status in cord blood is linked to adiposity in children between 3 and 7 years of age^23^, as well as up to 18 years of age^24^. Cord blood methylation profiles in children were shown to be influenced by maternal methylation profiles and by environmental factors that impact pregnancy^25,26^. Additionally, MRSs have been found to be associated with BMI in adults^27^, as well as in children^28^. However, the MRSs used in previous children studies were informed by BMI Epigenome Wide Association Studies (EWASs) in adults^28,29^, so it is still not known whether there are specific gene methylation patterns at birth that are linked to early childhood growth.

In this work, we capitalized on a cohort of second-born siblings to participants in the Intervention Nurses Start Infants Growing on Healthy Trajectories (INSIGHT) Study^6,8^. Specifically, these “SIBSIGHT” study participants were part of an observation-only longitudinal evaluation of second-born siblings. For this study, we investigated whether early childhood growth is associated with methylation in cord blood and placenta samples of 48 children from the SIBSIGHT cohort. Early childhood growth of children from the INSIGHT and SIBSIGHT cohorts has been extensively studied, providing evidence for a successful early-life intervention aimed at preventing childhood obesity for both siblings^7,8,30^. Along with insights into the effects of dietary intake^31,32^, sleep^33,34^, and infant temperament^35^, prior findings by our group have identified associations between early childhood growth and the composition of the oral microbiome^36^, the gut metabolome^37^, the stool micro-transcriptome^38^, and the genome^14^. Characterizing an association between gene methylation at birth and weight outcomes in children complements such studies, providing another avenue for identifying risk factors, adapting interventions–and thus preventing early life obesity and later life comorbidities. Here we used Illumina methylEpic arrays to establish genome-wide methylation profiles for placenta and cord blood tissues, and leveraged a wealth of additional information collected by SIBSIGHT. We tested a hypothesis that gene body methylation profiles at birth could be associated with weight outcomes in the first six months after birth.

## Results

We collected placenta and cord blood samples at the time of birth from 48 SIBSIGHT study participants^6,32^. For each sample, we used the Illumina MethylationEPIC array to determine methylation profiles across 575,132 CpG sites genome-wide. After quality control and clustering (see Methods for details), we grouped methylation signals from 293,090 CpGs into 20,108 genes. The number of CpG sites per gene ranged from 1 to 814, with average and median counts of 15 and 7, respectively.

We evaluated three weight outcomes of participating children: the conditional weight gain *z*-score (CWG, a standardized measure of change in weight from birth to six months of age, see Methods for details), BMI (weight/length^^2^) at six months, and the ratio of weight-for-length at six months. All three measures showed regular, Gaussian-like distributions across our participants (Figure S1; the Shapiro-Wilk test did not reject normality; CWG *p*-value = 0.416, BMI *p*-value = 0.529, weight-for-length *p*-value = 0.269). Weight outcomes at six months were chosen as they are the first outcomes we measured after collection of samples at birth; methylation patterns may change over time and could be modifiable^39,40^.

### Impact of covariates on weight outcomes

Prior to evaluating the associations between methylation profiles and weight outcomes, we assessed whether non-epigenetic covariates showed significant associations with the latter and should therefore be taken into account in downstream analyses. The non-epigenetic covariates we considered (Table 1) were maternal BMI and health-related variables (presence/absence of gestational diabetes, gestational weight gain, presence/absence of illness, and medication usage during pregnancy), gestational length, sex of the child, and infant feeding mode (i.e. breastfeeding or formula) at the age of 4, 16, and 28 weeks. Only one participating mother reported smoking, so this variable was excluded from the analysis. In order to determine which, if any, of the non-epigenetic covariates had an association with the infant weight outcomes, LASSO regressions were performed^41^ using each of the three weight outcomes as the response and the above-listed covariates as predictors. The only significant associations found were those of the sex of the child with weight-for-length and BMI (Figure S2). To account for this, we standardized these two weight outcomes by sex (see Methods for details). The CWG calculation already accounts for sex.

**Table 1.** Summary of SIBSIGHT Covariates used in the analysis. SD—standard deviation.

| Covariate | Value |
| --- | --- |
| Mother BMI<br>Average (SD) | 24.5 (4.5) |
| Father BMI<br>Average (SD) | 28.6 (4.5) |
| Child sex<br>N = female (%) | 27 (56%) |
| Gestational Duration (weeks)<br>Average (SD) | 38.9 (1.1) |
| Mode of Delivery<br>N = vaginal (%) | 34 (70.8%) |
| Maternal Age (years)<br>Average (SD) | 31.8 (4.3) |
| Gestational Diabetes<br>N = controlled by diet & exercise (%) | 3 (6.25%) |
| Smoking During Pregnancy<br>N = smoked | 1 |
| Maternal Illness during pregnancy (e.g.:<br>Thyroid disorders) (N=none) | 47 |
| Maternal Medications During Pregnancy<br>N = took medications (%) | 33 (68.8%) |
| Infant Feeding Mode at 4 weeks<br>N ≥ 80% breast milk (%) | 32 (66.7%) |
| Infant Feeding Mode at 16 weeks<br>N ≥ 80% breast milk (%) | 25 (52.1%) |
| Infant Feeding Mode at 28 weeks<br>N ≥ 80% breast milk (%) | 19 (39.6%) |

### Differences in methylation profiles between cord blood and placenta

The methylation state of a CpG site is determined by calculating the ratio between the methylated and unmethylated fluorescent signals from the microarray. This ratio is referred to as the methylation Beta signal^42^. The distribution of the methylation Beta signals across CpG sites differed between the two tissues analyzed (shown for each of the 48 children in Figure 1). The cord blood samples had the expected bimodal Beta value distribution with a strong peak at β<0.2 (hypomethylated CpGs) and a less pronounced peak at β>0.7 (hypermethylated CpGs). However, the placenta samples had a poorly defined peak at β>0.7, with more CpGs having values between β=0.2 to β=0.7. This suggests that our placenta samples contained either hemimethylated CpGs or heterogeneous cells with a mix of CpG methylation profiles.

**Figure 1.**
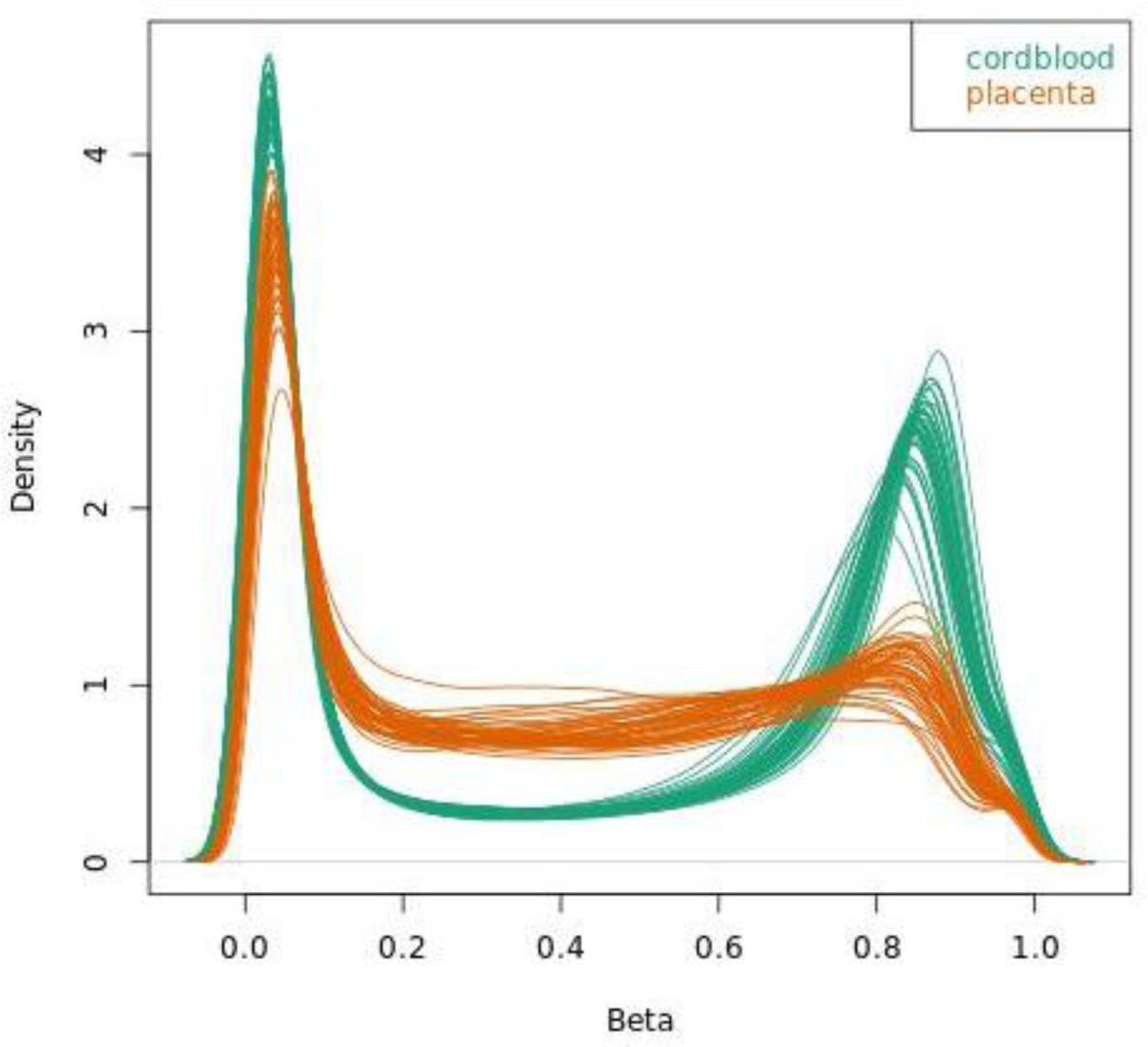
Density plots of Beta values describing the methylation state of CpG sites. Each line corresponds to an individual sample. Smoothing was performed with the function density plot from the Minfi package in R. The distributions for the 48 cord blood samples are shown in green, and those for the 48 placenta samples are shown in orange.

### Association study to identify differentially methylated genes

To identify genes with methylation patterns associated with children’s weight outcomes, we again used LASSO regression. In total, we performed six regressions, one for each tissue type and weight outcome combination. For each regression, we computed the average methylation states (Beta signals) across the CpGs for each gene, and used these averages as predictors. The results are summarized in Figure 2A. The LASSO regressions for cord blood and placenta identified, respectively, eight and ten genes whose methylation levels were significant predictors of CWG. Additionally, LASSO regressions identified four and 27 genes whose methylation levels in cord blood were significant predictors of BMI and weight-for-length, respectively (Table S4). In contrast, we did not identify any genes whose methylation in placenta was significantly associated with these two weight outcomes. Notably, in cord blood, there were three genes (*PLIN4*, *PPP1R16B,* and *UBE2F*) whose methylation levels were selected as significant predictors of all three weight outcomes, with similar coefficient estimates in the three regressions. There were no ‘shared genes’ among those selected for cord blood and placenta (Figure 2B). We report estimated coefficients from the LASSO regressions in Tables S3-S6; these express effect strength and sign: a positive regression coefficient can be interpreted as a higher methylation level being associated with an increased weight outcome, and a negative regression coefficient as a higher methylation level being associated with a decreased weight outcome.

**Figure 2.**
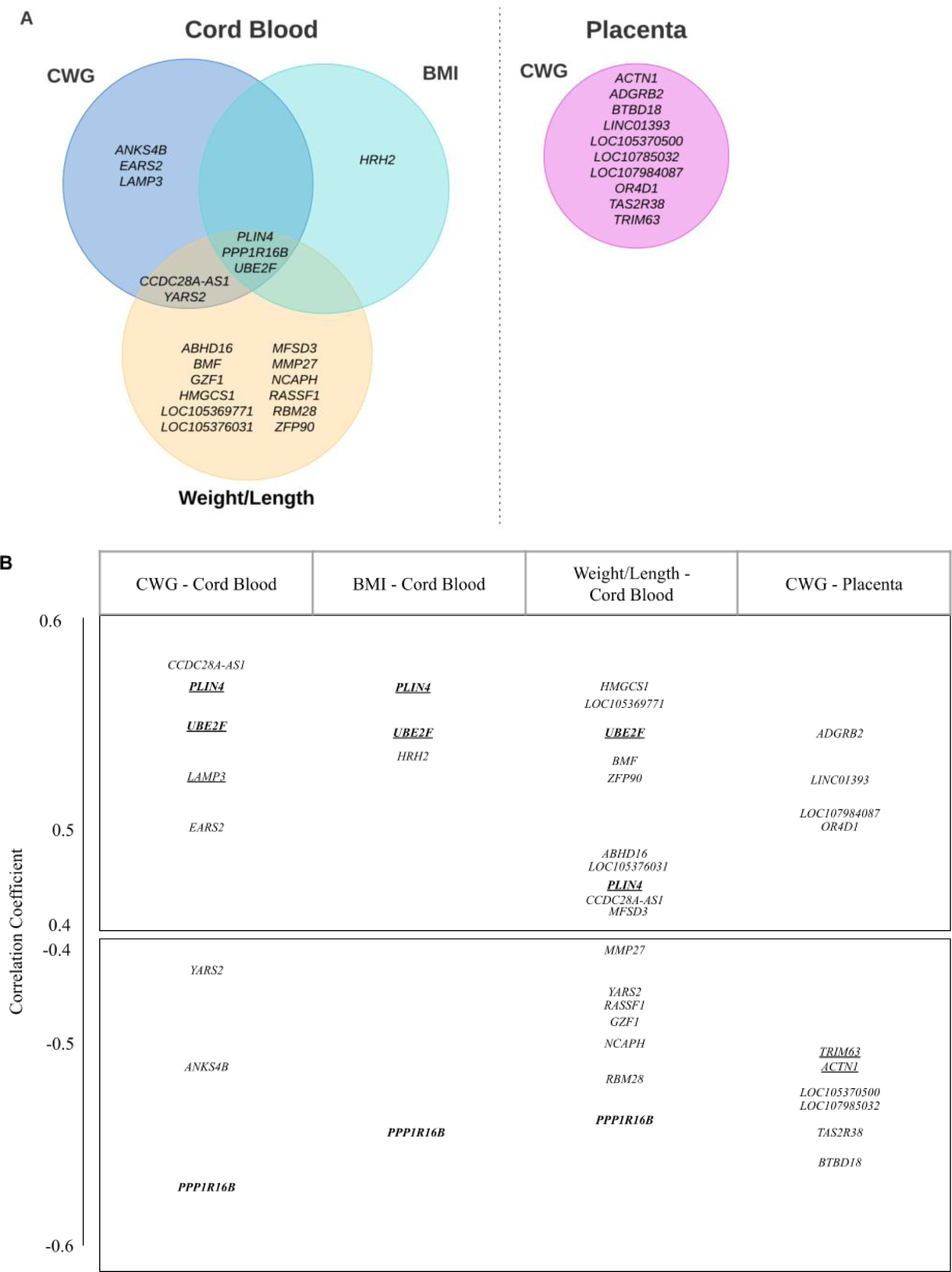
Genes whose methylation levels in cord blood and placenta are predictive of weight outcomes. The outcomes considered are conditional weight gain (CWG), body mass index (BMI), and weight-for-length (weight divided by length). **(A)** a Venn diagram of the relevant genes, as identified by LASSO regressions. **(B)** gene placement along the vertical axis corresponds to the correlation coefficient between each gene selected by the LASSO fit and the weight outcome. In bold are genes selected across multiple outcomes, and underlined are genes associated with weight outcomes in previous studies (see Discussion). Only CWG was associated with differentially methylated genes in the placenta.

We found that several genes selected in SIBSIGHT were also significantly related to child weight outcomes in an independent dataset—the PROGRESS^43^ cohort. PROGRESS is a freely accessible dataset of children from Mexico City, and comprises both cord blood DNA methylation data and longitudinal growth information for the children. Considering CWG as the weight outcome, and regressing it on one gene at a time, seven out of the eight genes selected in SIBSIGHT had a significant p-value also in PROGRESS. When regressing CWG on all eight genes jointly though, only *PPP1R16B* remained marginally significant (Table S7). Considering six-month BMI as the weight outcome, two out of four genes (*PLIN4* and *UBE2F*) selected in SIBSIGHT were significant in the joint regression in PROGRESS (Table S8). Finally, considering six-month weight-for-length as the weight outcome, one (*SMIM20*) of the 27 genes selected by SIBSIGHT was significant and one gene (*UBE2F*) was marginally significant in the joint regression (Table S9). It is notable that the genes that were selected using multiple weight outcomes in SIBSIGHT (*PLIN4, PPP1R16B,* and *UBE2F*) were also significantly associated with phenotypes in the independent PROGRESS cohort.

### Methylation Risk Score

Using results from the above LASSO regressions for the SIBSIGHT cohort, we generated a methylation risk score (MRS) for each growth outcome. These are weighted scores calculated as linear combinations of gene methylation Beta signals weighted by regression coefficient estimates obtained from post-LASSO Ordinary Least Squares fits (see Methods for details). Figure 3 shows the relationship between each MRS and the corresponding growth outcome. The associations were strong and significant in all cases, with high in-sample R-squared (cord blood CWG, adjusted R-squared = 0.874, *p*-value ≤ 2.2 x 10^-16^, Figure 3A; placenta CWG, adjusted R-squared = 0.8088, *p*-value ≤ 2.2 x 10^-16^, Figure 3B; cord blood BMI, adjusted R-squared = 0.992, *p*-value ≤ 2.2 x 10^-16^, Figure 3C; weight-for-length, adjusted R-squared = 0.5966, *p*-value 7.731 x 10^-11^, Figure 3D). Furthermore, there was still a significant relationship between these scores (calculated with phenotypes at 6 months) and the corresponding phenotypes at 1 and 2 years (Table S10). Using the independent PROGRESS cohort, however, these MRSs did not have a significant relationship with weight outcomes (Table S11).

**Figure 3.**
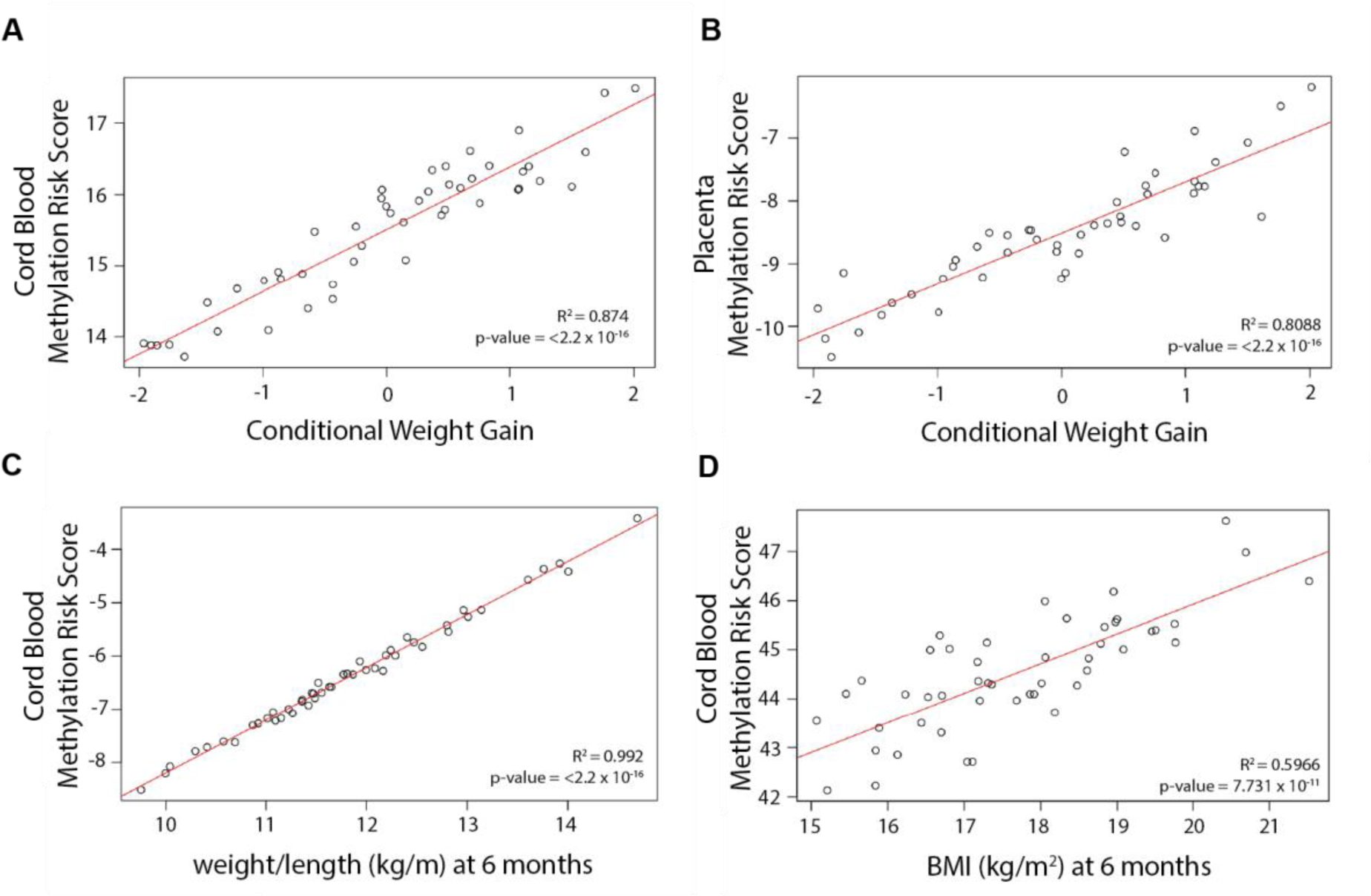
Relationship between MRS and weight outcomes. (A) Cord Blood MRS vs. Conditional Weight Gain. (B) Placenta MRS vs. Conditional Weight Gain. (C) Cord Blood MRS vs. weight-for-length ratio. (D) Cord Blood MRS vs. Body Mass Index. Note: Placental methylation does not produce a methylation risk score for BMI or weight-for-length as there was no relationship between gene methylation patterns and either of these weight outcomes.

## Discussion

In this study, we analyzed the methylation profiles of placenta and cord blood samples collected at birth. Using three outcomes characterizing early childhood growth, we identified genes whose methylation levels in these tissues are associated with weight gain during the first six months after birth. Comparing results of LASSO regression runs as well as the Ordinary Least Squares (OLS) regression of selected predictors across weight outcomes and tissues (Figure S5), we found that CWG and BMI provide more reliable results in cord blood when reproducing the analysis, with CWG having a higher adjusted R-squared than BMI (0.87 and 0.64 respectively). The weight-for-length ratio in cord blood had the highest R-squared (0.97) but the results were more variable. Indeed, attempts to replicate the correlation analysis with the weight-for-length ratio led to frequent low quality LASSO plots (with no associated genes or no clear minimum mean squared error), and a highly variable list of correlated genes, even after filtering for *p*-value (see Methods). For the placenta methylation, only CWG exhibited correlation with a set of gene methylation states, whereas BMI and weight-for-length did not. More generally, we found that, compared with the placenta methylation data, the cord blood methylation data presented a lower number of mixed methylation profiles, more genes associated with weight outcomes, and higher R-squared of the OLS regression on identified predictors (0.68 for predictors associated with CWG in the placenta).

### Genes whose methylation levels in cord blood are predictive of weight outcomes

In cord blood, we found three genes whose methylation levels were significantly associated with all three weight outcomes in SIBSIGHT and with outcomes in an independent cohort (PROGRESS). These are discussed below, followed by a discussion of genes identified as significant predictors for only one of the outcomes.

### PLIN4

One of the genes significantly associated with all three weight outcomes, and always with a positive sign (higher methylation inducing higher weight outcomes), was *PLIN4.* The protein encoded by this gene (Perilipin 4) is a member of the PAT family of lipid storage droplet proteins^44^. It is an important regulator of lipid storage. Low levels of expression of this protein have been associated with an increase in weight status^45^ of adults. Changes in *PLIN4* methylation have been observed after weight loss, with hypermethylation in the promoter region before vs. after gastric bypass surgery in adults^46^. *PLIN4* has also been classified as a putative obesogen in children, and was shown to be differentially methylated between obese and non-obese children in another study^47^.

### PPP1R16B

Another gene significantly associated with all three weight outcomes, and always with a negative sign (higher methylation inducing lower weight outcomes), was *PPP1R16B.* The protein encoded by *PPP1R16B* is phosphatase 1 (*PP1*) regulatory inhibitory subunit 16B^48^, which is also referred to as *TIMAP* or *ANKRD4*^49^. PP1 is involved in many essential cellular mechanisms and is part of a large interactome with over 200 interactors identified in vertebrates^50^. Studies of *PPP1R16B* showed its high levels of expression in endothelial cells and suggested that PP1 is involved in endothelium stability and permeability^49^. The activity of *PPP1R16B* has been shown to play a role in several diseases, including obesity and diabetes mellitus^49^.

### UBE2F

Finally, the third gene significantly associated with all three weight outcomes, and always with a positive sign (higher gene methylation inducing higher weight outcomes), was *UBE2F.* The protein encoded by *UBE2F* (Ubiquitin Conjugating Enzyme E2F) is a ubiquitin-protein ligase involved in post-translational modifications of proteins through the addition of ubiquitin-like protein NEDD8^51^. Previous studies have shown an association between the expression of this gene and BMI in children^52^. In animal models, *UBE2F* has been shown to be expressed at higher levels in the adipose tissue of obese rats compared to lean rats^53^.

### Other genes

We also identified several genes whose methylation level was significantly associated with only one weight outcome. Some such genes were also associated with obesity or an obesity-related trait in previous studies. One category of genes we identified were genes linked to nutrient metabolism, e.g. *ANKS4B*, *LAMP3*, as well as *PPP1R16B* (discussed above). The protein encoded by *ANKS4B* (Ankyrin Repeat And Sterile Alpha Motif Domain Containing 4B) plays a role in the epithelial brush border differentiation, controlling the microvilli organization and length^54^. It is involved in pancreas development and function^55,56^, affecting the secretion of insulin. This function could explain its link to weight gain. In our study, we found a negative association between CWG and cord blood methylation levels of *ANKS4B*. The protein encoded by *LAMP3* (Lysosomal Associated Membrane Protein 3) is involved in hepatic lipid metabolism and is overexpressed in patients with non-alcoholic fatty liver disease as well as in obese mice^57^. Our analysis indicated that *LAMP3* methylation is positively associated with CWG. The 33 additional genes implicated by our study but not already documented in the literature as being linked to obesity or metabolism (see Tables S3-S5) should be further analyzed in functional studies aimed at determining how they may influence weight gain in early childhood.

### Genes whose methylation levels in placenta are predictive of weight outcomes

In placenta, we found ten genes whose methylation levels were significantly associated with the CWG outcome (see Figure 3, Table S6). Four were identified as being involved in body weight and weight gain in prior studies, two have not been previously associated with obesity or obesity related traits in adults, and four are putative and of unknown function. Among previously studied genes, *TRIM63,* encoding for E3 ubiquitin ligase MURF1, has been linked to skeletal muscle atrophy and is over-expressed in obese rats compared to lean rats^58,59^. Methylation levels of *TRIM63* had a negative association with CWG in our study. *ADGRB2* is part of the adhesion G-protein-coupled receptor genes family, which is linked to insulin secretion in humans^60^ and modulation of adipogenesis and adipocyte function^61^. We found that methylation levels of *ADGRB2* had a positive association with CWG. *ACTN1* has been shown to be involved in adipogenesis^62^ and weight regain after weight loss^63,64^. In rats, it is up-regulated in the brain of animals with a high-fat diet^65^. *ACTN1* had a negative association with CWG in our study. Finally, *TAS2R38* has been shown to be involved in the perception of bitter taste^66^, and unrelated studies documented a link between the perception of bitterness and obesity in adults^67,68^ and male children^69^. Methylation levels of *TAS2R38* had a negative association with CWG in our study. These links suggest that these genes should be investigated further.

### Methylation risk scores as predictors of weight outcomes

A growing trend in genetics is to generate polygenic risk scores (PRS) for complex diseases because these types of disorders are often influenced by a large number of genetic variants, each with a small effect size^70^. PRSs, while not deterministic, can indicate which patients have a higher risk of developing certain conditions, which can aid in the establishment of intervention and/or treatment plans. MRSs have a similar advantage, capturing the cumulative effect of many CpG sites, or in this case the methylation signal of several genes, with small effect sizes. We developed MRSs for three phenotypes at six months after birth with cord blood methylation data from the SIBSIGHT cohort. Importantly, these MRSs remained significantly correlated with weight outcomes up to two years later.

### Conclusions and future directions

In this study we identified genes whose methylation levels in cord blood and placenta are significantly associated with three different weight outcomes; conditional weight gain *z*-score, BMI, and weight-for-length. Notably, we identified three genes whose methylation in cord blood is predictive of all three three weight outcomes. Two of these genes, *PLIN4* and *UBE2F*, have been associated with weight in prior studies. Also notably, and somewhat in contrast, only one outcome (CWG) was associated with gene methylation in the placenta. This can be explained by a higher number of cell types in the placenta tissue, making it more difficult to identify specific methylation patterns across a large number of methylation profiles. Alternatively, methylation states in the placenta might only be associated with CWG as birthweight is considered in the calculation of this outcome. It is possible that the conditions in the placenta might be more likely to influence birth weight than postnatal growth. One limitation of this study is the small sample size (48) compared to traditional Epigenome-Wide Association Studies (EWAS). In order to increase the power of our analysis, CpG sites were grouped by gene to reduce the dimensionality of the data, with the drawback that this allows us to capture only large-scale associations (i.e. over the whole gene and not individual CpGs). To confirm our findings, our analysis should be replicated using a larger sample.

We used the PROGRESS/ELEMENT DNA Methylation Study Dataset to test our selected genes and MRSs in an independent cohort. However, while the SIBSIGHT cohort is largely white and non-hispanic/latino^32^, the PROGRESS cohort is composed of individuals located in the Latin American city of Mexico City, Mexico^43^. It has been shown that in adults there is population-to-population variation in DNA methylation related to several diseases and phenotypes (e.g. cancer and diabetes)^71^ and that individuals who have similar demographics, life style, etc. have more similar methylation patterns^72^. Interestingly, we found evidence of between-populations differences in the association between weight outcomes and methylation patterns emerging as early as six months after birth. We found that the strongest “gene signals’’ from SIBSIGHT (*PLIN4, UBE2F,* and *PPP1R16B*) could also be detected in several of the regressions run on PROGRESS data. However, our MRSs were not predictive of weight outcomes in the PROGRESS cohort. We hypothesize that the underlying genetic, demographic, etc. differences between the two populations could be the reason why results from SIBSIGHT could not be more consistently validated in PROGRESS. This is notable because differences between the two cohorts were expected, however such a distinct contrast at such an early age was not. This suggests that external factors influencing the patterning of CpG methylation in early life should be carefully studied in order to determine factors potentially affecting future weight outcomes (e.g. maternal pre-pregnancy BMI^24^ or environmental exposures^43^). It will be beneficial to identify if there are shared patterns because these could be used to generate a MRS that could be used universally to identify the children most at risk for developing obesity and therefore benefit the most from targeted obesity prevention programs.

In this study we characterized methylation patterns within the gene body and not within the promoter regions^73^. The relationship between gene body methylation and gene expression has been shown to be U-shaped in some studies, with both high and low expression corresponding to high levels of methylation^74^, but in other studies methylation and transcription have been found to be positively correlated^20^. Additional studies are needed to fully investigate the expression levels of the gene bodies in both placenta and cord blood. Such studies could validate our findings and provide a better understanding of the mechanisms eventually affecting weight outcomes. To our knowledge, there are no gene body methylation studies investigating the large-effect obesity genes, e.g., *LEPTIN* and *FTO*, in infants. Notably, methylation of these two genes was not found to be associated with weight outcomes in our study.

In a prior study by our group^14^, we found that there may be different genetic components influencing infant weight gain vs. adult weight gain. Regulatory mechanisms, including methylation patterns, could therefore differ between adults and infants as well. This represents an interesting direction for future research; overall, methylation levels decrease throughout childhood and adolescence^75^ and it would be of great interest to investigate how the signatures we found here would persist as an individual ages.

## Methods

### Methylation Data Collection

We collected 48 matching samples of cord blood and placenta tissue from children enrolled in the SIBSIGHT study^6,31^. A list of the covariates employed in our analysis, with their summary values across the children included in this study can be found in Table 1.

At the time of birth, cord blood samples were collected in K_2_EDTA coated vacutainers (Becton, Dickinson, and Company) and stored at 4℃ until picked up by the research team. Samples were then stored at -80℃. DNA was isolated using the Qiagen DNeasy Blood and Tissue kit (Qiagen). Purified genomic DNA was then bisulfite-converted using EZ Methylation Kit (Zymo Research).

Placentas were stored at 4℃ after delivery before processing. 1cm^3^ pieces of the placenta were dissected from the fetal side, proximal to the area where the umbilical cord attaches. Tissues were formalin-fixed and paraffin-embedded. DNA from these tissues was extracted with the ReliaPrep FFPE gDNA Miniprep System (Promega) and then assessed with the FFPE QC kit (Illumina) for quality. Samples passing quality thresholds were then bisulfite-converted with the EZ Methylation Kit (Zymo Research), and then treated DNA was restored following the Infinium HD FFPE Restoration protocol (Illumina).

Bisulfite-converted DNA from both tissues was then analyzed on the Infinium MethylationEPIC chip (Illumina) in the Genome Sciences Facility at Penn State Hershey College of Medicine.

### Weight Outcomes Data Collection

For each child enrolled in this study, weight and length (via recumbent length board, Shorr Productions) were collected at birth and six months after birth, and BMI (kg/m^2^) and weight-for-length (kg/m) were calculated. Additionally, conditional weight gain (CWG) z-scores were calculated for each child using anthropometrics at birth and six months, adjusted for sex and age^76^. CWG *z*-scores are the standardized residuals from a linear regression of the weight-for-age *z*-score at six months on the weight-for-age *z*-score at birth (length-for-age *z-*score at birth and six months and exact age at the six-month visit are used as covariates in the regression). CWG *z*-scores are normally distributed and have a mean of 0 and a standard deviation of 1. Positive *z-*scores indicate above average weight gain (i.e., rapid infant weight gain) compared to other infants in the sample, and have been shown to be a risk factor for obesity later in life^77^. We standardized the BMI and weight-for-length ratio data by sex to remove the impact of the differences between sexes on the association with methylation profiles. This standardization is done by separating the two populations by sex and, for each, subtracting the mean and dividing by the standard deviation. Tests for normality were performed in R using the base stats package.

### Methylation Data Preprocessing

Raw signal reads from the chip were converted into Beta signals (β = intensity of the methylation signal/[intensity of the methylation signal + intensity of the unmethylated signal + 100]) using the Minfi package in R^78^. The Minfi package was also used to screen the data for quality. This included screening the data for outliers, excluding sex chromosomes, and excluding sites with known SNPs that could have caused false positives or negatives (see Table S1 for a summary of removed CpGs). After quality control, one placenta sample was removed from further analyses due to the low quality of the methylation data. Next, the Farray signals were normalized. First, we normalized within the array, which included background correction and normalization of signal intensity. Each chip contains control sites used to normalize between samples. Second, we utilized the Beta Mixture Quantile (BMIQ) normalization (one of the most popular methods found in the literature for MethylEpic analyses^79^) to normalize the signal from the Infinium I (InfI) and Infinium II (InfII) probes utilized on the MethylationEPIC array. BMIQ decomposes density profiles in three states: unmethylated, hemimethylated, and fully methylated. It rescales the InfII distribution to the corresponding InfI distribution. Both normalization steps were performed utilizing tools within the Minfi package (Figure S2).

After preprocessing we have one Beta signal for each CpG site, which corresponds to its methylation level. These values range from 0 (fully unmethylated) to 1 (fully methylated). Using the default density plot function included in the Minfi package, we visualize the distribution of individual CpG sites methylation levels. In a sample containing a single cell type, with identical methylation states between cells, we expect two peaks near 0 and 1. Values in between 0 and 1 indicate a mix of unmethylated and methylated sites in the sample, which can indicate a mix of cell types or cell states.

### Regression Analyses

To identify factors that could impact the weight outcomes (BMI) other than methylation profiles and sex, we performed a LASSO regression analysis^80,81^ (a method that performs predictor selection) on environmental factors, such as feeding mode, and family history, such as parental BMI and pregnancy duration (Table 1). We found no significant associations (Figure S2).

After the normalization performed during preprocessing, we grouped the methylation data by genes. Specifically, we averaged the Beta signals of CpG sites contained within the genomic coordinates of a gene to calculate the gene’s methylation level (Figure S3). We then ran LASSO regressions separately for the two tissues and, for each tissue, considering the three different weight outcomes–for a total of six regressions. We used the R package glmnet (LASSO and Elastic-Net Regularized Generalized Linear Models). The tuning parameters used for various LASSO runs can be found in Table S2; they were selected minimizing the cross-validation Mean Squared Error, as shown in the standard result plots produced by glmnet. Some of the LASSO fit analyses gave variable results for correlation with weight-to-length outcome. When repeating the analyses with the same parameters, the shape of the LASSO plot was changing, and, while a few genes were repeatedly selected, some results were not reproducible. To select the most predictive genes when the LASSO gave very variable results, we repeated the analysis until we obtained 10 profiles with the “check mark” shape plot, and selected the best model that included the most commonly selected genes across the replicate 10 analyses. After running each of the LASSO regressions, in order to reduce the bias, this technique creates in the estimation of the regression coefficients, we performed a post-selection fit–i.e. an OLS fit restricted to the set of predictors selected by the LASSO. We also ran marginal regressions for each individual predictor selected by the LASSO; the coefficient estimates from these regression can be considered alongside those produced by the post-selection OLS joint fit, as additional quantifications of the effects of each selected predictor.

### Methylation Risk Scores Calculation

Methylation risk scores (MRS) were calculated as described in ^22^. Briefly, they are a sum of *m* gene methylation values *c* (from section “Methylation data preprocessing”) with OLS estimated regression coefficients as weights *w* (from section “Regression analyses”):

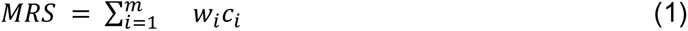

MRSs were calculated for each weight outcome separately. The association between MRS and weight outcome was determined by linear regression using the lm function in the basic stats package of R ^82^ using MRS as the predictor and weight outcome as the response.

### Validation datasets

Cord blood methylation data from the PROGRESS cohort^83^ was used in validation analyses (dbGaP: phs002754.v1.p1). This cohort consists of 1,001 individuals from Mexico City who were followed from birth through 18 years. Methylation data was downloaded from dbGaP (phs002754.v1.p1) and height and weight data were provided by study authors^83^. CWG z-scores, six-month BMI, and six-month weight-for-length were all calculated as described above.

To validate our results on the PROGRESS cohort, we preprocessed the data as we did for the SIBSIGHT cohort (see above), and confirmed the absence of association with the covariates available for this dataset (mother BMI, smoking, and disease during pregnancy). We performed linear regressions on the genes that were selected as predictors in the SIBSIGHT cohort. These regressions have been run both for each individual gene and as joint regression using all of the genes. We used the CWG *z*-scores to perform the linear regression with the genes associated with CWG in the SIBSIGHT cohort, and similarly for the BMI and weight/length ratio.

MRSs were calculated as described above, using the regression coefficients from SIBSIGHT as weights, *w.* As with SIBSIGHT, the association between the MRS and the phenotypes were calculated using the linear regression (lm) function in the basic stats package of R.

## Data Availability

All data produced in the present study are available upon reasonable request to the authors.

## Acknowledgements

We thank Bonnie Higgins and Kate Anthony for their assistance in sample processing and sample preparation, Dr. Ana Kenney and Dr. Debmalya Nandy for their statistical help, and the Penn State College of Medicine Genome Sciences Facility for generation of the methylation array data. We are also grateful for and thank the INSIGHT/SIBSIGHT families for their participation and the study nurses for their assistance in sample and data collection.

This study was supported by grants R01DK099354 and R01DK88244 from the National Institute of Diabetes and Digestive and Kidney Disease (NIDDK). The content is solely the responsibility of the authors and does not necessarily represent the official views of the NIH. Funding was also provided by Penn State Institute for Computational and Data Sciences, Penn State Eberly College of Sciences, and the Huck Institutes of Life Sciences at Penn State.

PROGRESS dataset: This work is supported by grants from the National Institute of Environmental Health Sciences (R01ES021357S1, R01ES021357, R01ES020268, R01ES013744). We acknowledge Instituto Nacional de Salud Pública, Instituto Nacional de Perinatologia, and the American British Cowdray (ABC) Hospital in Mexico City for their assistance with sample collection and use of their facilities. The PROGRESS/ELEMENT Mexico City cohort Principal Investigator is Robert Wright, Icahn School of Medicine at Mount Sinai.

