## Supplemental figures and tables for "Methylation profiles at birth linked to early childhood obesity"

### Table of Contents:

Supplementary Figure 1. Phenotype distribution 2
Supplementary Figure 2. BMI standardization LASSO result 2
Supplementary Figure 3. Preprocessing workflow 2
Supplementary Figure 4. Methylation analysis workflow 4
Supplementary Figure 5. Methylation LASSO results 4
Supplementary Table 1. Number of CpGs included at each step of the analyses 7
Supplementary Table 2. Parameters used for LASSO Analyses. 7
Supplementary Table 3. Genes identified in differential methylation analysis of conditional weight gain (CWG) in cord blood ordered from highest to lowest OLS correlation coefficients. 7
Supplementary Table 4. Genes identified in differential methylation analysis of Body Mass Index (BMI) in cord blood ordered from highest to lowest OLS correlation coefficients. 10
Supplementary Table 5. Genes identified in differential methylation analysis of weight/length ratio in cord blood ordered from highest to lowest OLS correlation coefficient. 10
Supplementary Table 6. Genes identified in differential methylation analysis of conditional weight gain (CWG) in placenta ordered from highest to lowest OLS correlation coefficient. 13
Supplemental Table 7. Regression coefficients and p-values from PROGRESS validation dataset usingConditional Weight Gain. 14
Supplementary Table 8. Regression coefficients and p-values from PROGRESS validation dataset using Body Mass Index. 14
Supplementary Table 9. Regression coefficients and p-values from PROGRESS validation dataset using Weight-for-Length. 15
Supplementary Table 10. Methylation Risk Score associations with phenotypes at later time points in the SIBSIGHT cohort. 17
Supplementary Table 11. Methylation Risk Score Associations with phenotypes in the PROGRESS cohort. 18

### Supplementary Figure 1. Phenotype distribution

**Distribution of (A)** CWG *z*-score from 0 to 6 months, (**B)** weight-for-length at six months, (**C)** BMI at the age of six months.

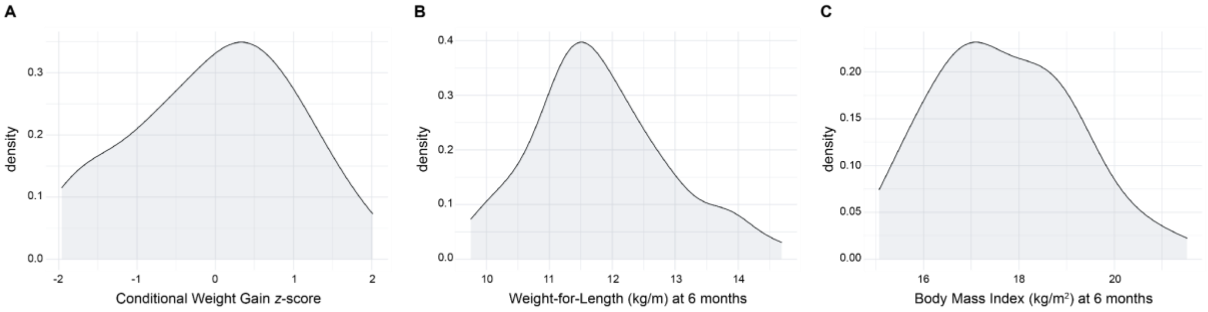

### Supplementary Figure 2. BMI standardization LASSO result

LASSO analysis to measure the association between covariates and BMI after standardization per sex. The absence of a stable minimum indicates no significant correlation between any other covariates and the phenotype.

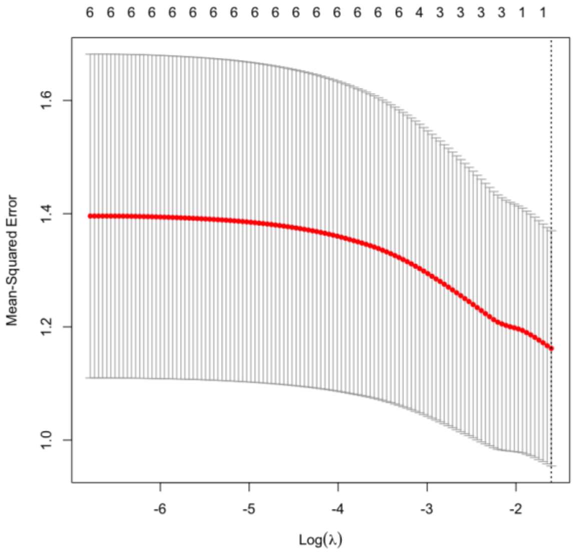

### Supplementary Figure 3. Preprocessing workflow

Workflow depicting the preprocessing of chip signal data. **(A)** Signal treatment and quality control. The raw data contain light intensity signals for each cell. The first step is to convert the light signal to methylation state of the corresponding CpG site, then to remove data that are of poor quality or that could artificially impact the association study such as sex chromosomes or probes with known SNPs. **(B)** Data normalization. Two types of normalizations are necessary. The first one normalizes the signal within each chip type. The second one normalizes between the two types of chips, allowing them to be used together in the analysis.

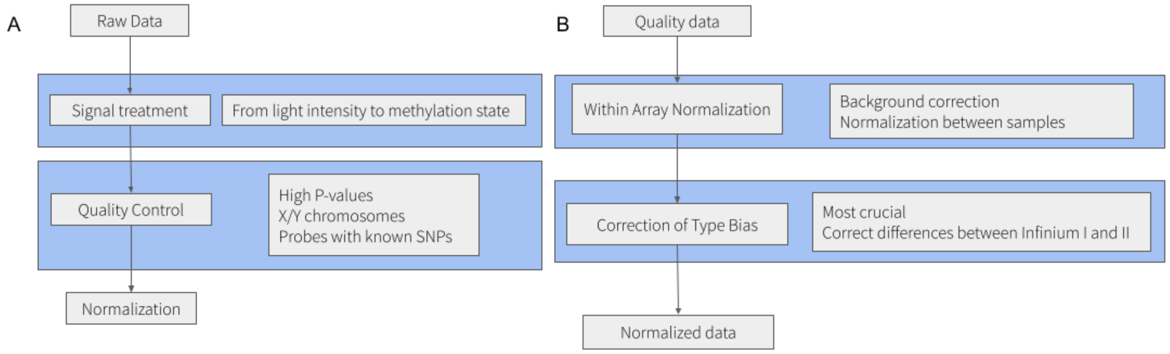

### Supplementary Figure 4. Methylation analysis workflow

Workflow depicting the association study to identify gene methylation profiles linked to weight outcome in children.

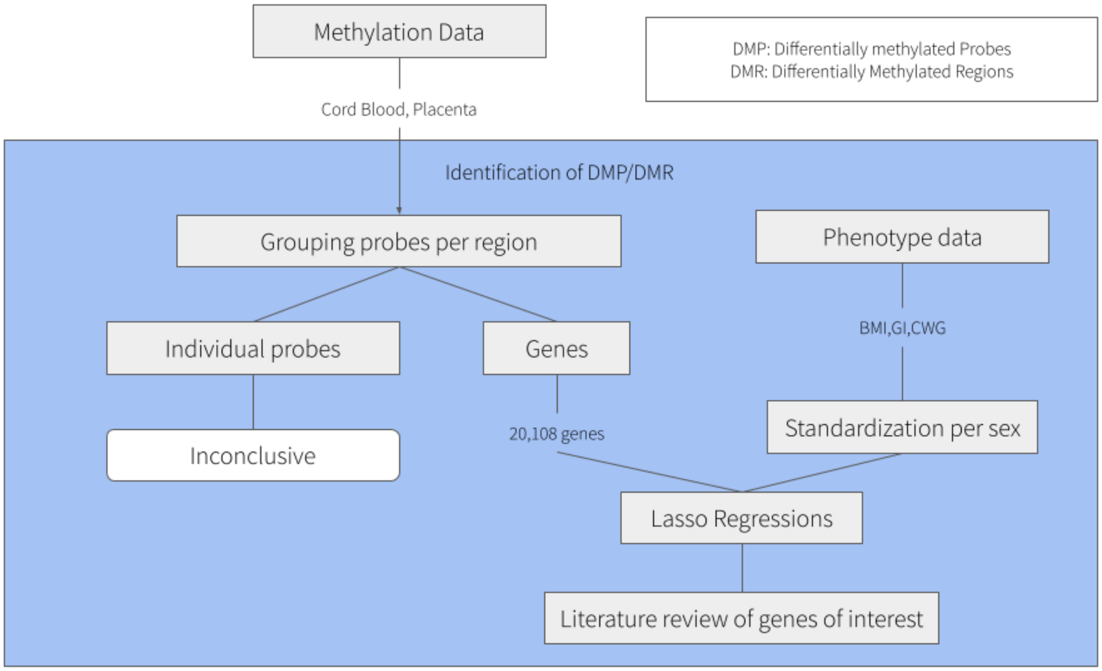

### Supplementary Figure 5. Methylation LASSO results

LASSO analysis to identify methylation profiles of genes linked to infant weight gain. (**A)** Methylated genes in cord blood associated with CWG. The LASSO analysis identified seven genes associated with CWG with a Mean-Squared error around 1.  (**B)** Methylated genes in the placenta associated with CWG. The LASSO analysis identified ten genes associated with CWG with a Mean-Squared error above 1.  (**C)** Methylated genes in cord blood associated with the BMI. The LASSO analysis identified four genes associated with CWG with a Mean-Squared error above 2.2.  (**D)** Methylated genes in placenta associated with the BMI. The LASSO analysis did not identify methylation to be associated with the BMI. (**E)** Methylated genes in the blood cord associated with the weight-for-length ratio. The LASSO analysis identified 27 gene methylations linked to the weight-for-length ratio, but the Mean-Squared error curve was almost flat and around 1.3. (**F)** Methylated genes in the placenta associated with the weight-for-length ratio. The LASSO analysis did not identify methylation to be associated with the weight-for-length ratio.

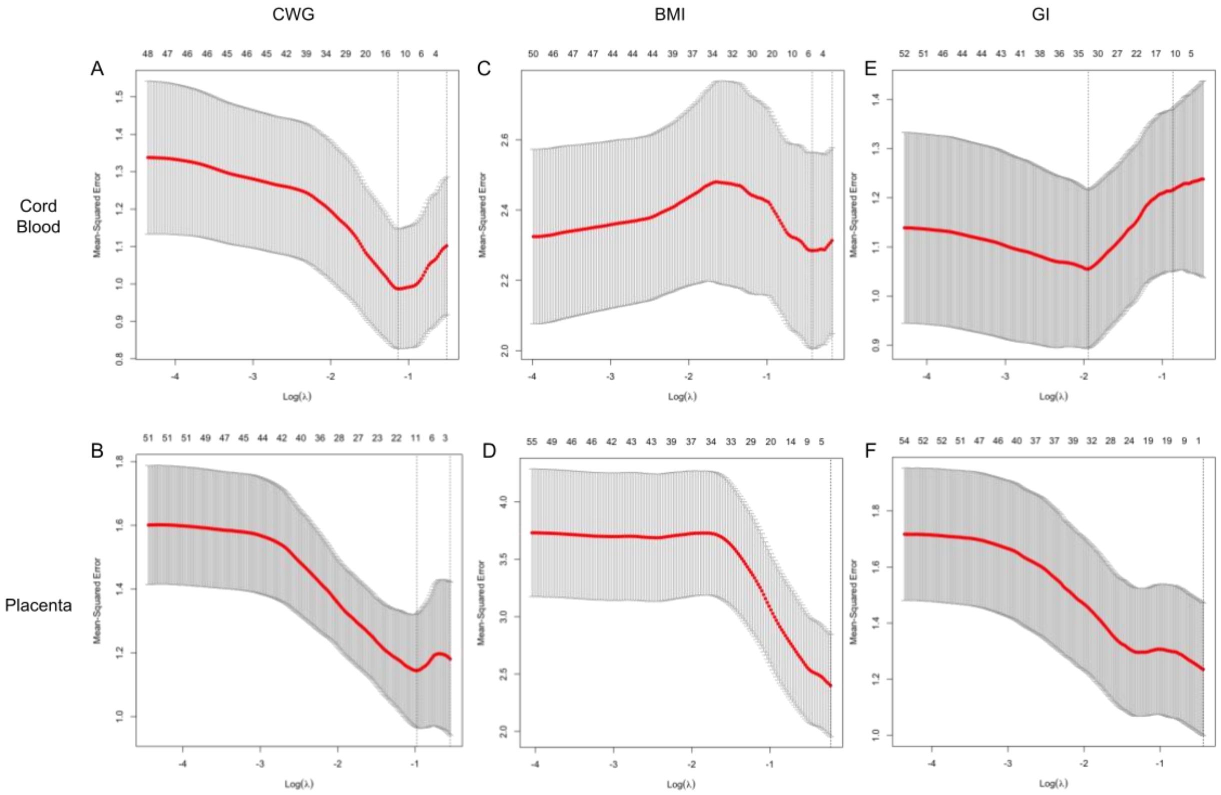

**Supplementary Table 1.** Number of CpGs included at each step of the analyses

| Tissue | Step | Number of CpGs remaining |
|---|---|---|
| Placenta | Initial | 575,132 |
| | Removing outliers and insufficient samples | 452,567 |
| | Removal of CpGs associated with common SNPs | 436,251 |
| | CpGs in genes | 293,090 |
| Cord Blood | Initial | 575,132 |
| | Removal outliers and insufficient samples | 452,567 |
| | Removal of CpGs associated with common SNPs | 436,251 |
| | CpGs in genes | 293,090 |

**Supplementary Table 2**. Parameters used for LASSO Analyses.

| Analysis | Nfold[1] | alpha[2] | nlambda[3] |
|---|---|---|---|
| Covariates | 10 | 1 | 200 |
| CWG Placenta | 10 | 1 | 300 |
| CWG Cord Blood | 10 | 1 | 200 |
| BMI Placenta | 10 | 1 | 200 |
| BMI Cord Blood | 10 | 1 | 200 |
| weight-for-length ratio Placenta | 10 | 1 | 300 |
| weight-for-length ratio Cord Blood | 10 | 1 | 300 |

[1] The number of folds to be used in the cross-validation
[2] Elastic net mixing parameter. alpha=1 is lasso regression (default) and alpha=0 is ridge regression
[3] The number of $\lambda$ values to be tested

**Supplementary Table 3.** Genes identified in differential methylation analysis of conditional weight gain (CWG) in cord blood ordered from highest to lowest Ordinary Least Squares(OLS) coefficients.

| Name | Chr | start | end | Function | Marginal regression Coefficient | Marginal p-value | Correlation Coefficient | OLS coefficient | OLS p-value |
|---|---|---|---|---|---|---|---|---|---|
| *LAMP3* | chr3 | 183122215 | 183162761 | lysosomal associated membrane protein 3 | 118.29 | 8.39E-05 | 0.54 | 28.84 | 8.61E-02 |
| *EARS2* | chr16 | 23520754 | 23557375 | glutamyl-tRNA synthetase 2%2C mitochondrial | 94.89 | 2.44E-04 | 0.51 | 23.63 | 1.22E-01 |
| *CCDC28A-AS1* | chr6 | 138725215 | 138773703 | CCDC28A antisense RNA 1 | 76.86 | 1.26E-05 | 0.58 | 23.49 | 3.29E-02 |
| *PLIN4* | chr19 | 4502192 | 4520285 | perilipin 4 | 45.45 | 1.33E-05 | 0.58 | 13.25 | 5.36E-02 |
| *UBE2F* | chr2 | 237966945 | 238042782 | ubiquitin conjugating enzyme E2 F (putative) | 93.30 | 3.07E-05 | 0.56 | 11.88 | 3.49E-01 |
| *STAP1* | chr4 | 67558727 | 67607337 | signal transducing adaptor family member 1 | 27.14 | 1.18E-03 | 0.45 | 9.33 | 4.82E-02 |
| *YARS2* | chr12 | 32725247 | 32756458 | tyrosyl-tRNA synthetase 2 | -12.91 | 9.17E-04 | -0.46 | -3.27 | 1.11E-01 |
| *PPP1R16B* | chr2 | 38805694 | 38923024 | protein phosphatase 1 regulatory subunit 16B | -71.25 | 1.71E-05 | -0.58 | -12.19 | 2.50E-01 |
| *TLK1* | chr2 | 170990823 | 171231293 | tousled like kinase 1 | -29.16 | 2.72E-03 | -0.42 | -15.59 | 2.52E-03 |
| *ANKS4B* | chr16 | 21233699 | 21253850 | ankyrin repeat and sterile alpha motif domain containing 4B | -27.62 | 1.55E-04 | -0.52 | -16.81 | 4.00E-05 |
| *LINC00486* | chr2 | 32825433 | 32946136 | long intergenic non-protein coding RNA 486 | -90.42 | 1.06E-03 | -0.46 | -24.12 | 9.89E-02 |

**Supplementary Table 4.** Genes identified in differential methylation analysis of Body Mass Index (BMI) in cord blood ordered from highest to lowest OLS coefficients.

| Name | Chr | start | end | Function | Marginal regression Coefficient | Marginal p-value | Correlation Coefficient | OLS coefficient | OLS p-value |
|---|---|---|---|---|---|---|---|---|---|
| *UBE2F* | chr2 | 237966945 | 238042782 | ubiquitin conjugating enzyme E2 F (putative) | 133.19 | 4.02E-05 | 0.56 | 45.07 | 1.14E-01 |
| *PLIN4* | chr19 | 4502192 | 4520285 | perilipin 4 | 64.05 | 2.41E-05 | 0.57 | 37.17 | 3.79E-03 |
| *HRH2* | chr5 | 175657762 | 175710756 | histamine receptor H2 | 54.76 | 6.58E-05 | 0.54 | 25.23 | 2.70E-02 |
| *PPP1R16B* | chr2 | 38805694 | 38923024 | protein phosphatase 1 regulatory subunit 16B | -98.27 | 4.88E-05 | -0.55 | -56.98 | 5.50E-03 |

**Supplementary Table 5.** Genes identified in differential methylation analysis of weight/length ratio in cord blood ordered from highest to lowest OLS coefficient.

| Name | Chr | start | end | Function | Marginal regression Coefficient | Marginal p-value | Correlation Coefficient | OLS coefficient | OLS p-value |
|---|---|---|---|---|---|---|---|---|---|
| *SMIM20* | chr4 | 25914192 | 25929879 | small integral membrane protein 20 | 96.04 | 2.76E-03 | 0.42 | 21.77 | 3.02E-02 |
| *ZFP90* | chr16 | 68530249 | 68576072 | ZFP90 zinc finger protein | 146.46 | 1.14E-04 | 0.53 | 19.83 | 8.83E-02 |
| *LOC105376031* | chr9 | 37027763 | 37031333 | | 65.46 | 5.27E-04 | 0.48 | 15.53 | 4.38E-03 |
| *ERP27* | chr12 | 14914027 | 14938537 | endoplasmic reticulum protein 27 | 56.14 | 1.36E-03 | 0.45 | 10.64 | 3.03E-02 |
| *MFSD3* | chr8 | 144508081 | 144511228 | major facilitator superfamily domain containing 3 | 108.96 | 8.27E-04 | 0.47 | 10.64 | 2.67E-01 |
| *HMGCS1* | chr5 | 43287470 | 43313477 | 3-hydroxy-3-methylglutaryl-CoA synthase 1 | 55.18 | 2.40E-05 | 0.57 | 9.31 | 3.49E-02 |
| *DNASE1* | chr16 | 3611737 | 3665472 | deoxyribonuclease 1 | 65.46 | 7.46E-03 | 0.38 | 9.26 | 1.98E-01 |
| *LOC105369771* | chr12 | 52105711 | 52119039 | | 47.54 | 3.34E-05 | 0.56 | 8.92 | 7.50E-03 |
| *BMF* | chr15 | 40087890 | 40108879 | Bcl2 modifying factor | 36.11 | 8.76E-05 | 0.54 | 7.06 | 2.33E-02 |
| *KCTD20* | chr6 | 36442767 | 36491143 | potassium channel tetramerization domain containing 20 | 15.05 | 1.99E-03 | 0.44 | 6.92 | 2.76E-05 |
| *PLIN4* | chr19 | 4502192 | 4520285 | perilipin 4 | 39.42 | 5.76E-04 | 0.48 | 2.86 | 5.25E-01 |

| | | | | | | | | | |
|---|---|---|---|---|---|---|---|---|---|
| *ABHD16A* | chr6 | 31686955 | 31703324 | abhydrolase domain containing 16A | 98.11 | 1.78E-04 | 0.52 | 2.47 | 7.48E-01 |
| *CCNG2* | chr4 | 77157207 | 77170060 | cyclin G2 | 14.56 | 3.20E-03 | 0.42 | 2.29 | 4.23E-02 |
| *CCDC28A-AS1* | chr6 | 138725215 | 138773703 | CCDC28A antisense RNA 1 | 65.40 | 7.38E-04 | 0.47 | 2.17 | 7.20E-01 |
| *GZF1* | chr2 | 23361585 | 23375399 | GDNF inducible zinc finger protein 1 | -11.06 | 3.01E-04 | -0.50 | -0.95 | 2.82E-01 |
| *YARS2* | chr12 | 32725247 | 32756458 | tyrosyl-tRNA synthetase 2 | -14.27 | 4.90E-04 | -0.48 | -1.54 | 2.24E-01 |
| *LOC107984670* | chr14 | 104741144 | 104741840 | | -24.62 | 3.96E-03 | -0.41 | -1.54 | 4.87E-01 |
| *LMTK2* | chr7 | 98106862 | 98209638 | lemur tyrosine kinase 2 | -24.29 | 1.08E-03 | -0.46 | -2.24 | 2.21E-01 |
| *FAM168B* | chr2 | 131047876 | 131093468 | family with sequence similarity 168 member B | -10.75 | 2.79E-03 | -0.42 | -2.30 | 2.73E-02 |
| *MMP27* | chr11 | 102690943 | 102705785 | matrix metallopeptidase 27 | -23.01 | 9.51E-04 | -0.46 | -3.71 | 1.29E-01 |
| *SLC6A2* | chr16 | 55655928 | 55706192 | solute carrier family 6 member 2 | -70.99 | 3.57E-03 | -0.41 | -4.95 | 4.01E-01 |
| *NCAPH* | chr2 | 96335766 | 96377091 | non-SMC condensin I complex subunit H | -95.21 | 2.28E-04 | -0.51 | -5.87 | 4.37E-01 |
| *RSF1* | chr11 | 77660009 | 77872232 | remodeling and spacing factor 1 | -117.51 | 3.05E-03 | -0.42 | -6.17 | 5.53E-01 |
| *RBM28* | chr7 | 128297685 | 128343915 | RNA binding motif protein 28 | -40.54 | 1.67E-04 | -0.52 | -10.03 | 1.04E-02 |
| *UBE2F* | chr2 | 237966945 | 238042782 | ubiquitin conjugating enzyme E2 F (putative) | 97.05 | 4.41E-05 | 0.55 | -11.81 | 1.14E-01 |
| *ABRAXAS1* | chr4 | 83459517 | 83485137 | abraxas 1%2C BRCA1 A complex subunit | -66.39 | 4.51E-03 | -0.40 | -12.80 | 5.43E-02 |
| *RASSF1* | chr3 | 50329786 | 50340936 | Ras association domain family member 1 | -143.51 | 4.60E-04 | -0.49 | -14.91 | 1.89E-01 |
| *PPP1R16B* | chr2 | 38805694 | 38923024 | protein phosphatase 1 regulatory subunit 16B | -70.34 | 7.68E-05 | -0.54 | -18.48 | 6.00E-04 |

| | | | | | | | | | |
|---|---|---|---|---|---|---|---|---|---|
| *LOC100129931* | chr4 | 7030554 | 7046231 | uncharacterized LOC100129931 | -70.10 | 6.01E-03 | -0.39 | -20.60 | 1.98E-02 |
| *LINC00486* | chr2 | 32825433 | 32946136 | long intergenic non-protein coding RNA 486 | -94.81 | 1.18E-03 | -0.45 | -20.83 | 2.10E-02 |
| *OBI1-AS1* | chr13 | 78054855 | 78617325 | OBI1 antisense RNA 1 | -164.46 | 2.67E-03 | -0.42 | -23.95 | 8.88E-02 |

**Supplementary Table 6.** Genes identified in differential methylation analysis of conditional weight gain (CWG) in placenta ordered from highest to lowest OLS coefficient.

| Name | Chr | start | end | Function | Correlation Coefficient | Marginal p-value | Marginal regression Coefficient | OLS coefficient | OLS p-value |
|---|---|---|---|---|---|---|---|---|---|
| *CUL4A* | chr13 | 113208193 | 113267108 | cullin 4A | 0.41 | 3.67E+01 | 0.00 | 15.80 | 6.17E-02 |
| *OR4D1* | chr17 | 58155154 | 58156086 | olfactory receptor family 4 subfamily D member 1 | 0.50 | 1.22E+01 | 0.00 | 3.60 | 1.11E-01 |
| *GDAP1L1* | chr2 | 44247099 | 44280937 | ganglioside induced differentiation associated protein 1 like 1 | 0.42 | 8.96E+00 | 0.00 | 2.90 | 1.52E-01 |
| *LOC107984087* | chr3 | 111783563 | 111798595 | | 0.50 | 9.29E+00 | 0.00 | 1.58 | 4.18E-01 |
| *TRIM63* | chr1 | 26051304 | 26067634 | tripartite motif containing 63 | -0.51 | -2.07E+01 | 0.00 | 1.45 | 7.40E-01 |
| *LINC01393* | chr7 | 115078958 | 115126314 | long intergenic non-protein coding RNA 1393 | 0.51 | 1.38E+01 | 0.00 | 0.46 | 8.82E-01 |
| *ADGRB2* | chr1 | 31727105 | 31764340 | adhesion G protein-coupled receptor B2 | 0.55 | 1.29E+01 | 0.00 | 0.10 | 9.69E-01 |
| *TAS2R38* | chr7 | 141972631 | 141973773 | taste 2 receptor member 38 | -0.56 | -6.55E+00 | 0.00 | -1.15 | 3.13E-01 |
| *LOC107985032* | chr17 | 35537124 | 35552311 | | -0.54 | -7.28E+00 | 0.00 | -2.88 | 4.46E-02 |
| *TM4SF19-TCTEX1D2* | chr3 | 196316085 | 196338420 | TM4SF19-TCTEX1D2 readthrough %28NMD candidate%29 | -0.46 | -9.87E+00 | 0.00 | -3.10 | 1.49E-01 |
| *BTBD18* | chr11 | 57743514 | 57753176 | BTB domain containing 18 | -0.56 | -9.11E+00 | 0.00 | -3.81 | 2.91E-02 |
| *LOC105370500* | chr14 | 52791777 | 52930234 | | -0.52 | -1.68E+01 | 0.00 | -3.99 | 2.06E-01 |

| | | | | | | | | | |
|---|---|---|---|---|---|---|---|---|---|
| *PHC1* | chr12 | 8914509 | 8941467 | polyhomeotic homolog 1 | -0.48 | -1.78E+01 | 0.00 | -4.99 | 1.77E-01 |
| *ACTN1* | chr14 | 68874123 | 68979366 | actinin alpha 1 | -0.52 | -2.74E+01 | 0.00 | -8.17 | 1.34E-01 |

### **Supplemental Table 7.** Regression coefficients and p-values from PROGRESS validation dataset using Conditional Weight Gain.

We used the genes selected in SIBSIGHT as predictors of CWG and performed a linear regression to evaluate their association with PROGRESS CWG scores.

| Name | coefcor[1] | coeflm[2] | pvalue[3] | jointcoeff[4] | jointpvalue[5] |
|---|---|---|---|---|---|
| *CCDC28A-AS1* | -0.18786 | -9.2411 | **0.0019341** | -6.1142 | 0.35063 |
| *EARS2* | -0.08657 | -5.7946 | 0.15603 | 3.4249 | 0.53584 |
| *LAMP3* | -0.15740 | -6.3765 | **0.0095817** | -2.75407 | 0.53975 |
| *LINC00486* | -0.16172 | -4.9535 | **0.0077556** | -0.64792 | 0.84838 |
| *PLIN4* | -0.12386 | -4.2955 | **0.041997** | 1.98906 | 0.66506 |
| *PPP1R16B* | -0.20240 | -16.2943 | **0.00082273** | -11.60958 | **0.051661** |
| *TLK1* | -0.12949 | -5.83659 | **0.033440** | 1.43766 | 0.74391 |
| *UBE2F* | -0.12657 | -6.0971 | **0.037673** | -2.13847 | 0.66633 |

[1] Correlation Coefficient
[2] OLS coefficient (Marginal)
[3] OLS pvalue (Marginal)
[4] OLS coefficient (Joint)
[5] OLS pvalue (Joint)

### **Supplementary Table 8.** Regression coefficients and p-values from PROGRESS validation dataset using Body Mass Index.

We used the genes selected in SIBSIGHT as predictors of BMI and performed a linear regression to evaluate their association with PROGRESS BMI scores.

| Name | coefcor[1] | coeflm[2] | pvalue[3] | jointcoeff[4] | jointpvalue[5] |
|---|---|---|---|---|---|
| *HRH2* | -0.013228 | -0.54713 | 0.82871 | -0.87206 | 0.80371 |
| *PLIN4* | -0.045939 | -1.0474 | 0.45220 | -5.2560 | **0.021123** |
| *PPP1R16B* | -0.036011 | -1.9059 | 0.55575 | -2.4344 | 0.460997 |
| *UBE2F* | 0.079208 | 2.5085 | 0.19445 | 8.7311 | **0.00401597** |

[1] Correlation Coefficient
[2] OLS coefficient (Marginal)
[3] OLS pvalue (Marginal)
[4] OLS coefficient (Joint)
[5] OLS pvalue (Joint)

### **Supplementary Table 9.** Regression coefficients and p-values from PROGRESS validation dataset using Weight-for-Length.

We used the genes selected in SIBSIGHT as predictors of weight-for-length ratio and performed a linear regression to evaluate their association with PROGRESS weight-for-length ratio scores.

| Name | coefcor[1] | coeflm[2] | pvalue[3] | jointcoeff[4] | jointpvalue[5] |
|---|---|---|---|---|---|
| *ABHD16A* | -0.06395 | -4.1153 | 0.29511 | 4.7511 | 0.57617 |
| *CCDC28A-AS1* | -0.039994 | -1.8159 | 0.51286 | 9.1543 | 0.20502 |
| *CCNG2* | -0.05653 | -1.02607 | 0.35480 | 0.73173 | 0.70032 |
| *DNASE1* | -0.085808 | -4.1959 | 0.15972 | -2.4685 | 0.60966 |
| *ERP27* | -0.064821 | -0.97326 | 0.28856 | -0.59163 | 0.73663 |
| *FAM168B* | 0.023452 | 0.47367 | 0.70125 | 0.640296 | 0.64617 |
| *KCTD20* | 0.020412 | 1.4286 | 0.73847 | 4.1064 | 0.49032 |
| *LINC00486* | -0.063162 | -1.7857 | 0.30110 | -1.5726 | 0.64533 |
| *LMTK2* | 0.0037663 | 0.22581 | 0.95088 | 2.6310 | 0.59831 |
| *LOC100129931* | 0.044005 | 4.7732 | 0.47149 | 14.808 | 0.11446 |
| *NCAPH* | -0.05556 | -1.15995 | 0.36312 | -1.6378 | 0.23809 |
| *OBI1-AS1* | -0.075565 | -7.8813 | 0.21584 | -5.9929 | 0.61726 |
| *PLIN4* | -0.044307 | -1.4183 | 0.46844 | -6.7985 | 0.17025 |
| *PPP1R16B* | -0.076829 | -5.7089 | 0.20823 | -0.85031 | 0.93693 |
| *RASSF1* | -0.086701 | -5.0332 | 0.15540 | -5.8135 | 0.379667 |
| *RBM28* | -0.062905 | -3.29198 | 0.30308 | -3.2668 | 0.63397 |
| *RSF1* | -0.0046131 | -0.21401 | 0.939861 | -3.9159 | 0.44889 |
| *SLC6A2* | -0.067418 | -3.1597 | 0.26963 | -7.9535 | 0.20460 |
| *SMIM20* | -0.063760 | -4.1448 | 0.29654 | -13.016 | **0.042614** |
| *UBE2F* | 0.039684 | 1.7645 | 0.51614 | 9.3191 | **0.066160** |

[1] Correlation Coefficient

[2] OLS coefficient (Marginal)
[3] OLS pvalue (Marginal)
[4] OLS coefficient (Joint)
[5] OLS pvalue (Joint)

### **Supplementary Table 10**. Methylation Risk Score associations with phenotypes at later time points in the SIBSIGHT cohort.

Methylation risk scores are calculated with the phenotype at 6 months. These are the linear regression coefficients of the relationship of those scores with phenotypes later in life.

| | Methylation Risk Score | Phenotype | Adjusted $R^2$ | p-value |
|---|---|---|---|---|
| Cord Blood | BMI | BMI @ 1year | 0.4189 | $1.726 \times 10^{-7}$ |
| | BMI | BMI @ 2years | 0.2503 | $2.03 \times 10^{-4}$ |
| | weight-for-length | Weight-for-length @ 1year | 0.5825 | $1.726 \times 10^{-10}$ |
| | weight-for-length | Weight-for-length @ 2years | 0.3716 | $3.247 \times 10^{-6}$ |
| | CWG | BMI @ 1year | 0.5431 | $1.415 \times 10^{-9}$ |
| | CWG | BMI @ 2years | 0.249 | $2.2115 \times 10^{-4}$ |
| | CWG | Weight-for-length @ 1year | 0.4125 | $5.185 \times 10^{-7}$ |
| | CWG | Weight-for-length @ 2years | 0.2401 | $2.8 \times 10^{-4}$ |
| Placenta | CWG | BMI @ 1year | 0.4571 | $1.107 \times 10^{-7}$ |
| | CWG | BMI @ 2years | 0.1891 | $1.483 \times 10^{-3}$ |
| | CWG | Weight-for-length @ 1year | 0.3359 | $1.177 \times 10^{-5}$ |

| | | | | |
|---|---|---|---|---|
| | CWG | Weight-for-length @ 2years | 0.183 | $1.774 \times 10^{-3}$ |

### Supplementary Table 11. Methylation Risk Score Associations with phenotypes in the PROGRESS cohort.

Methylation risk scores were derived from the SIBSIGHT cohort. These scores were then calculated using methylation data from the PROGRESS cohort. The data shown here are the results of linear regressions of the MRS and phenotypes.

| | Methylation Risk Score | Phenotype | Adjusted $R^2$ | p-value |
|---|---|---|---|---|
| Cord Blood | CWG | CWG | -0.002118 | 0.5108 |
| | | BMI @ 1 year | -0.001999 | 0.4744 |
| | | BMI @ 1.5 years | -0.004094 | 0.9081 |
| | | BMI @ 2 years | -0.002947 | 0.599 |
| | | Weight/Length @ 1 year | -0.004087 | 0.9338 |
| | | Weight/Length @ 1.5 years | -0.004038 | 0.8701 |
| | | Weight/Length @ 2 years | -0.001793 | 0.4551 |
| | BMI | BMI @ 6 months | -0.001946 | 0.4919 |
| | | BMI @ 1 year | -0.00314 | 0.6274 |
| | | BMI @ 1.5 years | -0.003981 | 0.8406 |
| | | BMI @ 2 years | -0.00374 | 0.7731 |
| | Weight/Length | Weight/Length @ 6 months | -0.001928 | 0.4897 |
| | | Weight/Length @ 1 year | -0.004101 | 0.9526 |

| | | | | |
|---|---|---|---|---|
| | | Weight/Length @ 1.5 years | -0.004077 | 0.895 |
| | | Weight/Length @ 2 years | -0.002835 | 0.5815 |
